# Analytical and clinical validation of a targeted-enhanced whole genome sequencing-based comprehensive genomic profiling test

**DOI:** 10.1101/2023.12.18.23300049

**Authors:** Stephanie Ferguson, Shruthi Sriram, Jonathan Kyle Wallace, Jeonghoon Lee, Jung-Ah Kim, Yoonsuh Lee, Brian Baek-Lok Oh, Won Chul Lee, Sangmoon Lee, Erin Connolly-Strong

## Abstract

Evaluation of the test performance of the targeted enhanced whole-genome sequencing (TE-WGS) assay for comprehensive oncology genomic profiling. The analytical validation of the assay included sensitivity and specificity for single nucleotide variants (SNVs), insertions/deletions (indels), and structural variants (SVs), revealing a revealed a sensitivity of 99.8% for SNVs and 99.2% for indels. The PPV was 99.3% for SNVs and 98.7% for indels. Clinical validation was benchmarked against established orthogonal methods and demonstrated high concordance with reference methods in variant characterization. The TE-WGS assay enhances personalized cancer treatment by offering detailed genomic insights and the adaptability to include emerging biomarkers.

## 1. Introduction

Clinical application of next-generation sequencing (NGS) is transforming the management of diseases by offering precise genetic insights for personalized therapy. High-throughput NGS aims to pinpoint unique mutational landscapes in tumors to inform tailored treatment strategies and/or enrollment in specific clinical trials. Empirical evidence supports the premise that precision medicine — which utilizes genomic and molecular profiling to tailor treatments based on specific tumor alterations — can significantly improve patient outcomes in terms of survival, quality of life, and economic efficiency when compared to traditional single-gene testing methods. [1,2].

In clinical settings, NGS assays typically focus on a selected array of pivotal genes, implement cancer-type-specific targeting techniques, or are limited to whole-exome sequencing, which concentrates exclusively on the genome’s coding regions[3]. Contrastingly, Whole Genome Sequencing (WGS) provides a comprehensive, unselected analysis of the entire genome, encompassing both the protein-coding and non-coding regions. This exhaustive approach has the potential to uncover a wider spectrum of actionable therapeutic targets [4], thereby broadening the scope for personalized treatment options.

WGS stands out from targeted panel or exome sequencing by eliminating biases associated with sequence capture, thus providing a more complete picture of the genomic landscape [5]. A critical point of consideration is the elevated false-positive rates often associated with tumor-only testing methods, a challenge notably pronounced in patients with non-European ancestry. This issue is compounded in standard tumor analysis practices, which typically involve referencing public Single Nucleotide Polymorphism (SNP) databases for filtering benign germline variants. Predominantly representing European genetic information, these databases exhibit limited efficacy for diverse ethnic populations [6,7]. Recent evidence suggests that relying solely on population databases to filter germline variations can lead to an overestimation of tumor mutational burden (TMB)[8]. Such overestimations could have significant implications, potentially skewing clinical trial results and impacting patient outcomes, especially in the realm of FDA-approved immunotherapies."

This report details the analytical and clinical validation of the CancerVision assay, a system employing targeted enhanced whole genome sequencing (TE-WGS) for comprehensive analysis of solid tumors. The report presents on the assay’s reproducibility, sensitivity, and detection thresholds for single nucleotide variants (SNVs), copy number variations (CNV), and structural variations. The unique methodology of CancerVision integrates TE-WGS with concurrent tumor and germline analysis, enabling the accurate characterization of mutational landscapes and structural variations within the tumor, as well as germline contributions. By effectively differentiating between tumor-specific alterations and inherited sequences, the assay significantly increases the accuracy of somatic variant detection across diverse populations. The assay’s robust performance is augmented by its integration with a database of FDA-approved treatments and clinical trials, enhancing the decision-making process for targeted therapies. By enabling the identification of both established and novel genomic alterations, CancerVision aims to refine the landscape of genomic profiling in cancer treatment, tailoring therapeutic approaches to the intricate genomic profile of each patient’s tumor.

## 2. Data and Methods

### 2.1 Test principle and intended use

The Targeted Enhanced Whole Genome Sequencing (TE-WGS) assay (CancerVision - Genome Insight, San Diego, CA), offers an integrated approach for genomic profiling by combining comprehensive whole genome analysis with a focused examination of targeted biomarker genes (refer to **Supplementary Table 1** for the gene list). This assay is adept at systematically identifying a wide range of genomic aberrations in solid tumor specimens. It detects single nucleotide variants (SNVs), multiple nucleotide variants (MNVs), small insertions and deletions (indels), copy number variations (CNCs), and structural variations (SVs). In addition to these detections, TE-WGS generates a detailed report highlighting the mutational landscape, including mutational signatures, and genomic instability status on: tumor mutational burden (TMB), microsatellite instability (MSI), and homologous recombination deficiency (HRD). The intended use of TE-WGS is to identify genomic alterations that have established and/or potential clinical relevance in the context of solid tumor malignancies.

### 2.2 Patient and Reference Materials

In this study, a dual-methodology approach was implemented, combining well-characterized reference materials with patient-derived specimens for robust analysis. The reference materials included cell lines from the Genome in a Bottle (GIAB) project and commercially obtained FFPE specimens, and DNA isolated from FFPE specimens, sourced from Horizon Discovery, United Kingdom, and the American Type Culture Collection, Manassas, VA. Specifically, the NA12878, NA24694, NA24695, NA24631, NA24143 cell lines from Genome in a Bottle (National Institute of Standards and Technology, US) were employed to assess the analytical performance of the study. Additionally, reference samples from Horizon Discovery, specifically the OncoSpan FFPE line, were selected for their representation of a diverse range of histopathological conditions and included both single nucleotide variants (SNVs) and insertions-deletions (indels) with previously validated allele frequencies.

The clinical validation of the TE-WGS assay was conducted using a select cohort of 56 residual patient samples. These samples were obtained from a prospective cohort registry study at Ajou University Medical Center. Selection was based on the availability of adequate residual formalin-fixed paraffin-embedded (FFPE) tissue, blood, and/or DNA for comprehensive analysis. The inclusion criteria for sample selection included a confirmed prior cancer diagnosis and patient age of 18 years or older at the time of sample collection. The study’s protocol was approved by the Ajou University Medical Center Independent Review Board, ensuring compliance with the ethical standards outlined in the Declaration of Helsinki, and all participants provided written informed consent.

For the purpose of this study, each cancer patient’s tumor sample comprised a minimum of five unstained slides at 5-micron thickness, alongside a corresponding hematoxylin and eosin (H&E) stained slide for tumor tissue delineation. A board-certified pathologist conducted a histopathological evaluation to ascertain the neoplastic content within these tissues.

For assay precision and quality control, the NA12878 cell line from the Coriell Institute in Camden, NJ, was utilized. This cell line known for its extensively documented human genome reference, providing a reliable external quality control measure throughout the validation process.

### 2.3 Regulatory standards

TE-WGS is conducted at Genome Insight, a facility accredited by the College of American Pathologists (CAP) and certified under the Clinical Laboratory Improvement Amendments (CLIA) in the United States. CLIA provides federal regulatory standards that apply to all clinical laboratory testing performed on humans in the United States, excluding clinical trials and basic research.

### 2.4 Library Preparation

For this study, DNA libraries were constructed using the Watchmaker DNA Library Preparation Kit (Watchmaker Genomics, Boulder, CO). The procedure began with the enzymatic fragmentation of DNA, followed by adapter ligation. Post-ligation, a bead-based cleanup process was executed, and subsequent library amplification was carried out. Key quality control measures included assessing the library size distribution targeting an average fragment size of approximately 300 bp, and quantifying yield, with a minimum threshold of 15 ng/uL. Library size was determined by the TapeStation 4200 System (Agilent Technologies, Santa Clara, CA). Concentration determinations were made using the Qubit DNA Assay Kit in conjunction with a Qubit 2.0 Fluorometer (ThermoFisher Scientific, Waltham, MA, USA). Prepared libraries were preserved at ≤ –20°C when not immediately processed to the capture stage, adhering to the kit manufacturer’s storage guidelines.

### 2.5 Sequencing and Data Analysis Pipeline

The comprehensive genomic analysis was performed using the TE-WGS CancerVision system (Genome Insight Inc., San Diego, CA, USA). Sequencing of the prepared DNA libraries was carried out on the Illumina NovaSeq 6000 sequencing system (Illumina Inc.), achieving an average target depth of coverage of 40x for tumor samples and 20x for matched blood samples for whole-genome sequencing (WGS). Target-enhanced sequencing for tumor DNA was facilitated by xGen Custom Hybridization Probes (IDT, Inc., Coralville, IA, USA), covering a genomic region of 2.76 Mb, and sequenced to an average target depth of 500x.

The resultant raw sequences were mapped to the human reference genome build GRCh38 using the BWA-MEM algorithm. PCR duplicate reads were excised using SAMBLASTER [9]. Variant calling for germline small variants utilized HaplotypeCaller and Strelka2[10,11], while somatic small variant detection employed Strelka2 and Mutect2 [10,11]. Structural variant identification was conducted via Manta[12]. All variants, both germline and somatic, were annotated using the Variant Effect Predictor (VEP) [13] and subjected to rigorous manual review and curation within Genome Insight’s proprietary genome browser.

Tumor purity, ploidy, and allele-specific copy number were determined using whole-genome sequencing (WGS) data. These parameters were inferred based on the depth ratio of tumor to normal data and B allele frequency. The primary tool used for these calculations was Sequenza, with supplementary steps implemented to correct tumor purity for copy number stable tumors and to mitigate noise in formalin-fixed paraffin-embedded (FFPE) specimens.

Final FASTQ files, Variant Call Format (VCF) files, and Compressed Reference-oriented Alignment Map (CRAM) files were securely packaged, encrypted, and transferred to a long-term storage solution. This process was designed to ensure both data integrity and confidentiality.

## 3 Results

### 3.1. Analytical Sensitivity and Positive Predictive Value for SNV and Indel Detection of Whole Genome Sequencing

A true-positive (TP) was a mutation of interest (MOI) in the commercial cell lines identified by both the GIAB consortium and the WGS assay. Conversely, a false-negative (FN) was a mutation recognized by the GIAB consortium but missed by the WGS assay. The assay’s sensitivity was quantified using the formula: TP / (TP + FN) = Assay Sensitivity. This calculation, applied to data from six reference cell lines (Genome in a Bottle), resulted in an analytical sensitivity of 99.8% for SNVs (95% CI: 99.79% - 99.83%) and 99.2% for indels (95% CI: 99.14% - 99.72%)—refer to **Table 1**.

**Table 1:** Overall performance of targeted enhanced-whole genome sequencing (TE-WGS)

|  | Variant | Specification |
| --- | --- | --- |
| **Analytical Sensitivity** | SNV | 99.8% (CI 99.79-99.83) |
|  | Indel | 99.2% (CI 99.14-99.72) |
| **Analytical PPV** | SNV | 99.3% (CI 99.28-99.38) |
|  | Indel | 98.7% (CI 98.63-98.78) |

Indel, insertion, and deletion; PPV, positive predictive value; SNV, single nucleotide variant

In this study, false positives (FP) were defined as mutations detected by the Whole Genome Sequencing (WGS) assay at specific loci, which were established as wild-type in commercial cell lines by the Genome in a Bottle (GIAB) consortium. To calculate the assay’s Positive Predictive Value (PPV), the formula: PPV = TP / (TP + FP) was used, where TP represents true positives. The TE-WGS analytical PPV, evaluated against the GIAB standard revealed a PPV of 99.3% for Single Nucleotide Variants (SNVs) (95% Confidence Interval [CI]: 99.28% - 99.38%) and 98.7% for insertions and deletions (indels) (95% CI: 98.63% - 98.78%), as detailed in Table 1.

The CancerVision Targeted Enhanced Whole Genome Sequencing (TE-WGS) test was subjected to analytical validation, which included assessing various parameters such as nucleic acid extraction, sequencing accuracy, and data analysis precision. The assay’s performance was evaluated using a spectrum of tumor-derived cell lines and reference standards from established commercial sources, alongside clinical formalin-fixed paraffin-embedded (FFPE) samples verified through orthogonal testing methods. TE-WGS achieved an average mean depth of coverage of 55.3x (ranging from 42.8x to 67.1x) for Whole Genome Sequencing (WGS), and 810.6x (ranging from 650.6x to 1147.4x) for Targeted Enhanced (TE) targeted panel sequencing (TPS).

The reference FFPE DNA samples, provided by Horizon, included SNVs and indels with previously validated allelic frequencies. The reference sample comprised 25 established variants. Excluding two variants (KIT D816V and PIK3CA E545K) with potentially lower sequencing efficiency due to vector sequences, comparative analysis of 23 variants between expected and detected allelic frequencies via the TE-WGS assay revealed strong agreement, with a correlation coefficient R=0.99 for SNVs and R=0.98 for indels (**Figure 1A and B**).

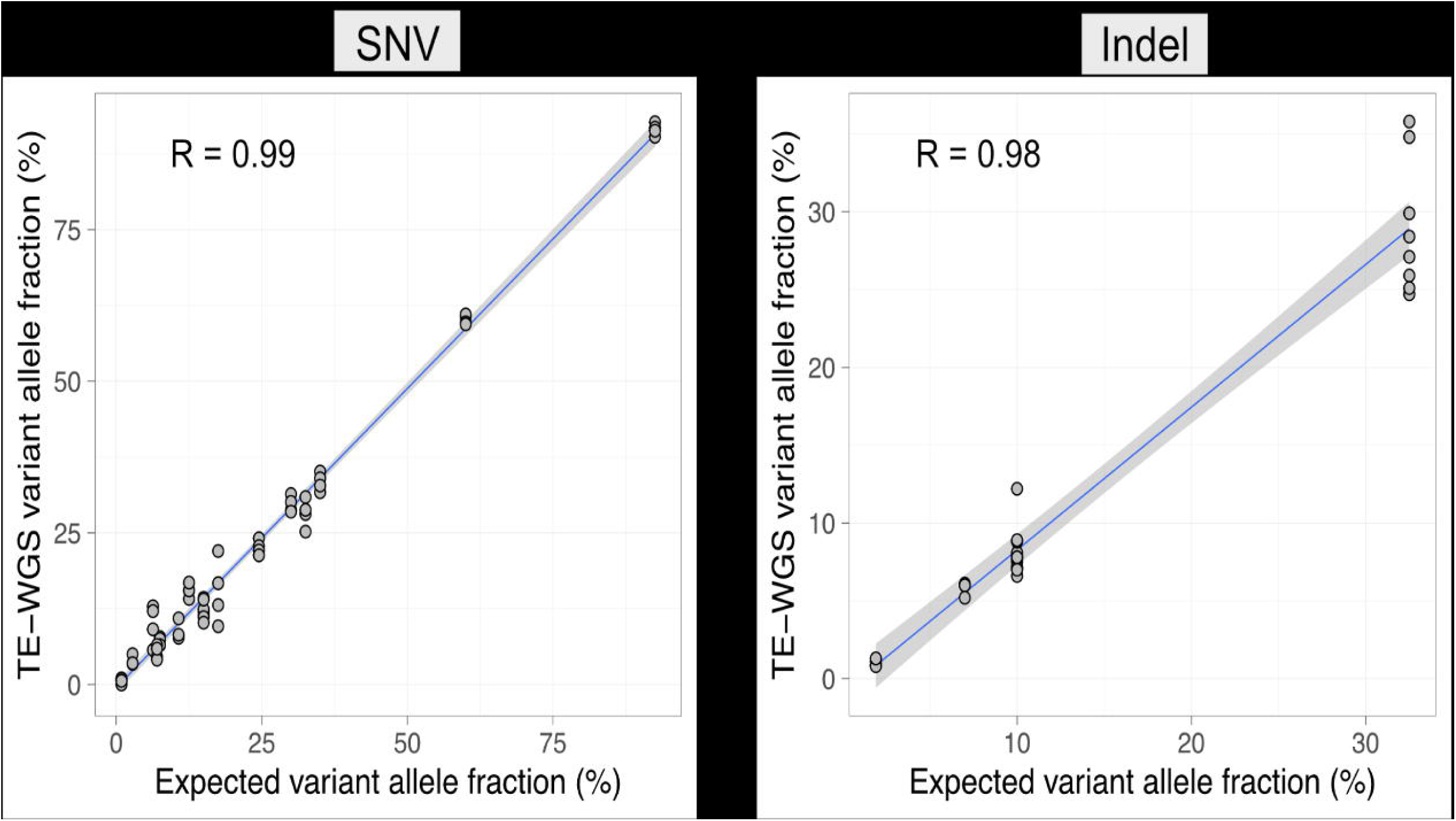

### 3.2 Limit of Detection (LOD)

#### Single Nucleotide Variants (SNVs)

The LOD for SNVs within the TE-WGS assay was assessed using a serial dilution approach. A KRAS G12V Reference Standard from Horizon, initially at a 50% variant allele frequency (VAF), was methodically diluted to generate VAFs of 1%, 5%, 10%, and 15%. NA12878 DNA, which harbors no KRAS variants, served as the diluent. As demonstrated in **Figure 2**, the TE-WGS assay’s detection of the KRAS G12V variant was consistent and linearly proportional to the expected VAFs. The minimum LOD for SNVs, based on these evaluations, has been established at a VAF of 5%.

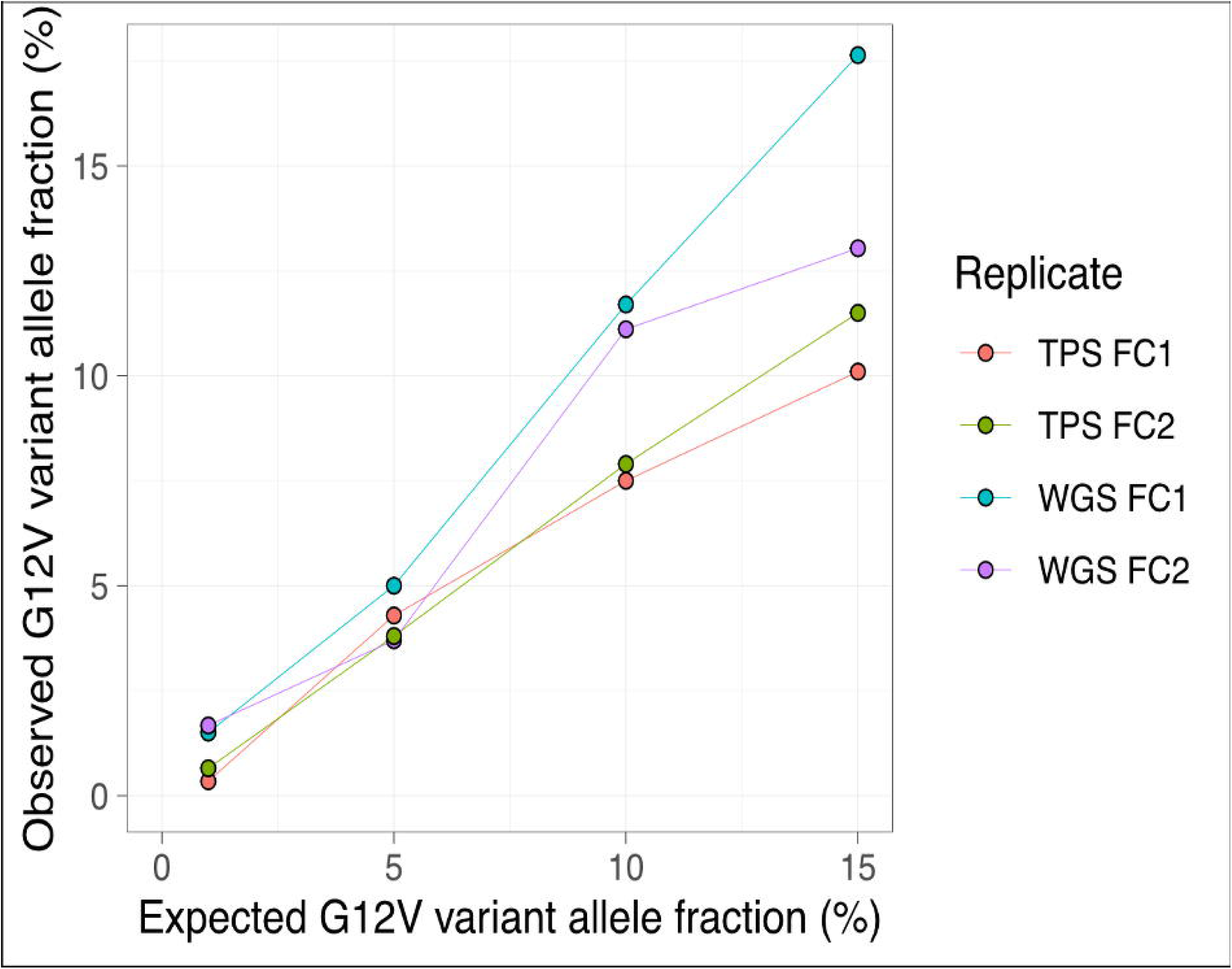

#### Insertions and Deletions (Indels)

For indels, the LOD was established by analyzing variant frequencies in OncoSpan FFPE samples from Horizon. The defined minimum LOD for indel detection stands at a VAF of 10%.

#### Structural Variations (SVs)

The LOD for SVs was determined using the Structural Multiplex Reference standard (FFPE curl) from Horizon, which encompasses a variety of known structural variations at distinct VAFs. From this analysis, the assay’s LOD for SVs has been set at a minimum VAF of 15%.

The above-stated LODs reflect the assay’s robustness in detecting variants of different types and sizes at low allelic frequencies, as substantiated by the comprehensive analytical validation data.

### 3.3 Reproducibility

For a comprehensive assessment, genomic DNA (gDNA) from each specimen was divided into three aliquots: two allocated for intra-laboratory testing by separate operators and one reserved for external orthogonal validation. This approach enabled parallel processing and thorough examination of reproducibility. The methodology yielded an inter-precision reproducibility rate of 95.8% (69/72 variants), demonstrating the reliability and robustness of the TE-WGS workflow.

Furthermore, the inter-assay precision was ascertained by comparing concordance between results from the TE-WGS assay and orthogonal testing. Of the 72 variants assessable by both platforms, the TE-WGS assay and orthogonal testing both failed to identify the same two variants, resulting in an overall inter-assay precision rate of 97.2% (70/72 variants). This finding further substantiates the accuracy and consistency of the TE-WGS assay relative to orthogonal testing methodologies.

### 3.4 Clinical Validation

Clinical validation of the assay encompassed an analysis of 28 paired tumor and normal clinical samples, yielding a cohort of 56 specimens (Table 2). The concordance of somatic variant identification was assessed by comparing the variant calling outcomes from our assay against those derived from a parallel orthogonal clinically validated sequencing assay, conducted by a CLIA-certified laboratory.

**Table 2:** Tumor types of the samples for clinical validation.

| Tumor types | Number of samples (percent) |
| --- | --- |
| Breast cancer | 12 (42.9%) |
| Stomach cancer | 6 (21.4%) |
| Head and neck cancer | 3 (10.7%) |
| Bladder cancer | 2 (7.1%) |
| Gallbladder cancer | 2 (7.1%) |
| Lung cancer | 1 (3.6%) |
| Colorectal cancer | 1 (3.6%) |
| Small bowel cancer | 1 (3.6%) |
| Total | 28 (100%) |

In the context of Copy Number Variations (CNVs), both the TE-WGS assay and Orthogonal Testing examined 15 CNVs. The TE-WGS assay missed the identification of 1 CNV, which resulted in a CNV inter-assay precision of 14/15, or 93.3%. Notably, the TE-WGS assay identified 64 CNVs not detected by Orthogonal Testing.

For Single Nucleotide Variants (SNVs), the comparison covered 57 SNVs. The TE-WGS assay did not detect one SNV, leading to an SNV inter-assay precision of 56/57, or 98.3%. Moreover, the TE-WGS assay identified 103 additional SNVs that were not reported by Orthogonal Testing.

We estimate microsatellite instability (MSI) by genome-widely examining microsatellite regions. The score reports the number of somatic insertions and deletions per Mb in microsatellite regions across the whole genome of the tumor. A tumor is considered microsatellite stable (MSS) if the score is < 20, and MSI-High if > 20. To validate the MSI test we performed accuracy studies using 28 patient samples tested by an orthogonal approach. Patient samples were selected to demonstrate a range of tumor content from 23% up to 80% (mean 46%) by tumor estimate. MSI status was all stable by TE-WGS, of which concordance rate with the orthogonal test was 100% (28/28).

The Tumor Mutation Burden (TMB) is quantitatively determined using the targeted enhanced whole-genome sequencing (TE-WGS) method, which incorporates a germline subtraction technique to ascertain the number of somatic changes within the entire genome (∼2.6Gb). A TMB value ≥ 10 mutations/Mb is classified as ‘high’. Comparison of TMB between two replicates of TE-WGS showed excellent correlation (R = 0.99), which implied the consistency of the TMB calculation by TE-WGS. Comparisons of TMB assessments between TE-WGS and an alternative tumor-only NGS methodology that uses an extensive panel revealed a notable correlation coefficient (R=0.81 in the replicate 1 and R=0.80 in the replicate 2). Despite the general agreement, there were discrepancies; specifically, 3 cases (10.7%) that the orthogonal testing classified as ‘high’ TMB were identified as ‘low’ TMB (<10 mutations/Mb) in the TE-WGS analysis.

### 3.5 Clinical Utility

The potential clinical utility of the TE-WGS assay was evaluated by quantifying the incidence of clinically actionable genetic alterations identified within the patient cohort. Clinically actionable alterations are characterized as those associated with FDA-approved therapeutics for on- or off-label use, as well as pathogenic or likely pathogenic germline variations. Within the study group, twelve patients (42.9%) harbored genomic aberrations for which there are commercially available therapies, excluding investigational treatments (accessible only via clinical trials). Notably, six patients (21.4%) presented with genomic modifications that were concordant with FDA-approved on-label pharmacological interventions specific to their cancer type. Moreover, one patient (3.6%) exhibited germline alterations deemed pathogenic or likely pathogenic according to established clinical criteria. Notably, two patients (7.1%) harbored complex rearrangements in the actionable genes, such as *BRIP1*, *ATR*, and *RAD51B*, which were challenging to detect with a targeted panel approach.

## 4 Discussion

In the evolving landscape of precision oncology, the utility of comprehensive genomic profiling (CGP) to direct targeted therapies and inform prognosis is well-established [14,15]. The CancerVision targeted enhanced whole-genome sequencing (TE-WGS) assay has been developed and analytically validated to support this paradigm by facilitating a nuanced examination of the genomic alterations that drive tumor behavior. The TE-WGS assay has undergone rigorous analytical and clinical validation, proving to be a reliable and accurate tool for comprehensive genomic profiling in oncology. The assay demonstrates high sensitivity and specificity for single nucleotide variants (SNVs), insertions and deletions (indels), and structural variations (SVs). The assay exhibits high sensitivity and specificity for single nucleotide variants (SNVs), insertions and deletions (indels), and structural variations (SVs). The validated limits of detection of the TE-WGS assay are particularly noteworthy, enabling the identification of clinically significant mutations at low allelic frequencies. This level of sensitivity is critical in detecting mutations that may be present in only a subset of tumor cells but could potentially drive therapeutic resistance.

The clinical validation of the TE-WGS assay has demonstrated its high accuracy in the detection of somatic variants, yielding results consistent with other established methods. In contrast to fixed-panel NGS assays, which are limited to known genomic alterations and can quickly become obsolete as new biomarkers emerge, the comprehensive nature of the TE-WGS assay allows for the capture of a wider spectrum of clinically relevant mutations. This attribute of the TE-WGS assay ensures that patients have access to the latest genomic discoveries without the delays associated with the continual updating and validation of new testing panels [3,16].

While newer comprehensive genomic tests may utilize tumor/germline comparison, test such as whole exome sequencing do not achieve the sequencing coverage of the TE-WGS assay. This disparity in sequencing coverage may lead to a reduction in accuracy and sensitivity when identifying clinically significant variants. Such limitations are particularly evident in Tumor Mutational Burden (TMB) analysis, where tumor-only assays or panel tests may overestimate TMB[8]. The precise quantification of TMB is crucial, as it has become an important biomarker for the eligibility of patients for immunotherapy, a rapidly advancing field in cancer treatment. The recent FDA approval of pembrolizumab for high TMB tumors exemplifies the clinical relevance of accurate TMB assessment, despite the ongoing need for more standardized measurement and reporting practices in this area[17]. While this study showed a good correlation between NGS panel test and TE-WGS approaches, instances of discordance were observed, indicating that TE-WGS methodology may have the potential to reduce exposure to ineffective therapeutic approaches.

## 5 Conclusion

The comprehensive results of this study demonstrate that the TE-WGS is a robust and reliable assay that accurately and reproducibly detects a patient’s genomic landscape. These data support the validity of TE-WGS in the clinical decision-making process of solid tumor patients. With a focus on adaptability to new oncological markers and a 14-day turnaround time, it is equipped to support a personalized cancer management approach.

## Data Availability

All data produced in the present study are available upon reasonable request to the authors

## Acknowledgments

The authors would like to thank all patients and Dr. Minsuk Kwon at Ajou University who participated in the clinical study.

## Funding

This research was supported by the Bio & Medical Technology Development Program of the National Research Foundation (NRF) funded by the Korean government (MSIT) (No. 2022M3A9G101451221) along with Genome Insight Inc.

## Conflict of Interest

The authors are employees of Genome Insight

## Ethics approval and consent to participate

This work combined data from the parent studies (IRB numbers: AJOUIRB-SMP-2022-278 and AJOUIRB-SMP-2021-576) that had previously obtained informed consent from participants and IRB approval. The authors state that they have followed the principles outlined in the Declaration of Helsinki.

## Consent for Publication

In alignment with ethical guidelines, written informed consent for the publication of their data and/or images was obtained from all participants involved in the study. This consent was part of the initial ethical approval process (IRB numbers: AJOUIRB-SMP-2022-278 and AJOUIRB-SMP-2021-576). Additionally, all patient information presented in this publication has been anonymized to protect patient privacy and confidentiality.

## Availability of Data

The patients participating in this study did not consent to the public release of sequencing data. The TE-WGS pipeline and associated algorithms are proprietary to Genome Insight Inc.

---

**Supplementary Table 1.**
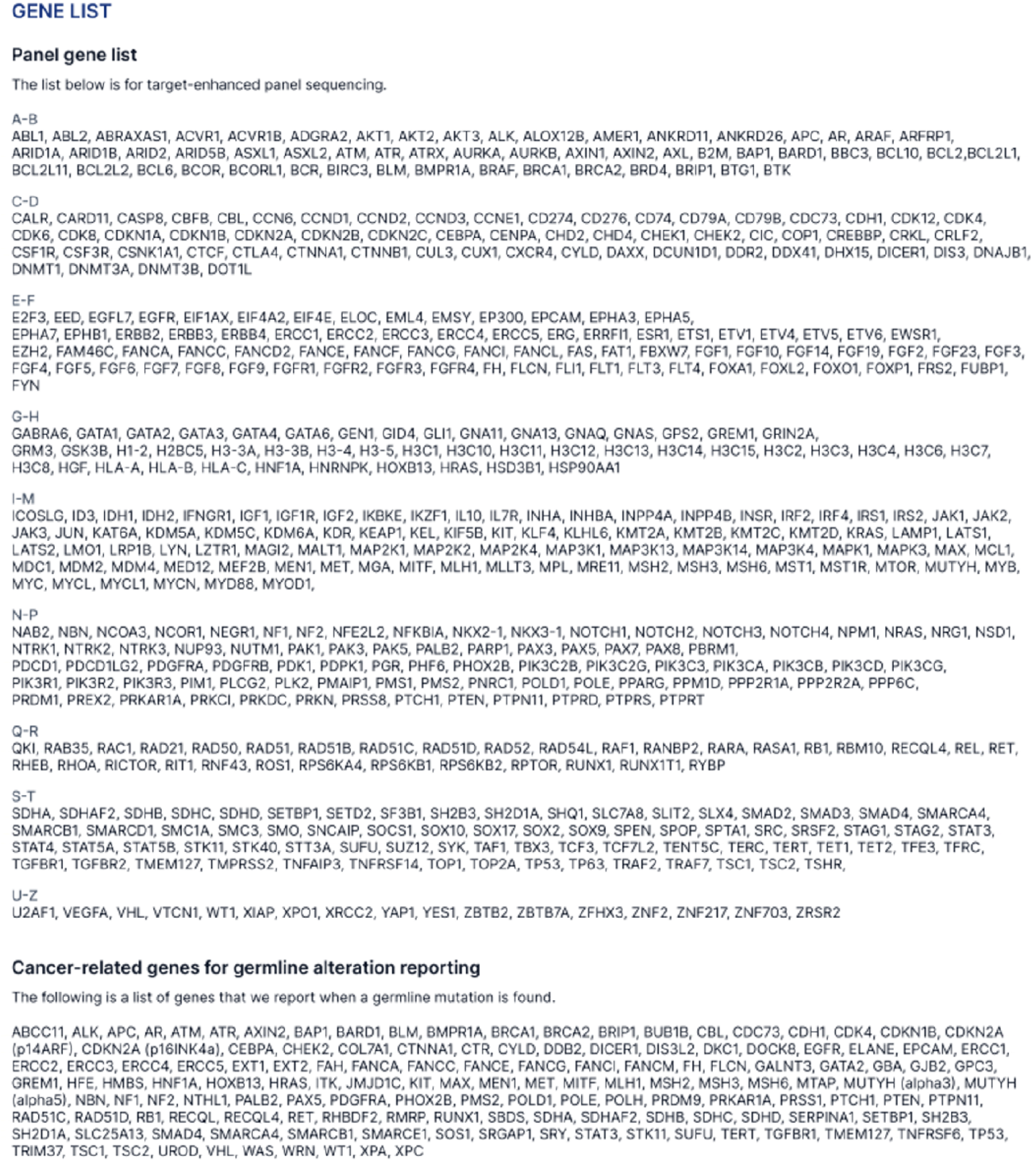

## Notes

### Competing Interest Statement

The authors of the publication are employees of Inocras Inc.

### Author Declarations

Clinical validation of the TE-WGS assay was performed using a selected cohort of residual patient samples (N = 56) obtained from a prospective cohort registry study (Ajou University Medical Center), based on the availability of adequate residual formalin-fixed paraffin-embedded (FFPE) material, blood, and/or DNA for comprehensive analysis. The chosen samples met the following inclusion criteria: a confirmed prior cancer diagnosis and patient age of ≥18 years at the time of sample collection. The study protocol received approval from the Ajou University Medical Center Independent Review Board, with all participants providing written informed consent in line with ethical standards set forth in the Declaration of Helsinki.

### Summary of Updates

To correct some text and table errors.

## References

[1] Leroy K, Audigier Valette C, Alexandre J, et al. Retrospective analysis of real-world data to evaluate actionability of a comprehensive molecular profiling panel in solid tumor tissue samples (REALM study). PLoS One. 2023;18(9):e0291495.

[2] Agarwala V, Khozin S, Singal G, et al. Real-World Evidence In Support Of Precision Medicine: Clinico-Genomic Cancer Data As A Case Study. Health Aff (Millwood). 2018 May;37(5):765–772.

[3] Malone ER, Oliva M, Sabatini PJB, et al. Molecular profiling for precision cancer therapies. Genome Med. 2020 Jan 14;12(1):8.

[4] Schwartzberg L, Kim ES, Liu D, Schrag D. Precision Oncology: Who, How, What, When, and When Not? Am Soc Clin Oncol Educ Book. 2017;37:160–169.

[5] Vijay P, McIntyre AB, Mason CE, et al. Clinical Genomics: Challenges and Opportunities. Crit Rev Eukaryot Gene Expr. 2016;26(2):97–113.

[6] Garofalo A, Sholl L, Reardon B, et al. The impact of tumor profiling approaches and genomic data strategies for cancer precision medicine. Genome Med. 2016 Jul 26;8(1):79.

[7] Halperin RF, Carpten JD, Manojlovic Z, et al. A method to reduce ancestry related germline false positives in tumor only somatic variant calling. BMC Med Genomics. 2017 Oct 19;10(1):61.

[8] Parikh K, Huether R, White K, et al. Tumor Mutational Burden From Tumor-Only Sequencing Compared With Germline Subtraction From Paired Tumor and Normal Specimens. JAMA Netw Open. 2020 Feb 5;3(2):e200202.

[9] Faust GG, Hall IM. SAMBLASTER: fast duplicate marking and structural variant read extraction. Bioinformatics. 2014 Sep 1;30(17):2503–5.

[10] McKenna A, Hanna M, Banks E, et al. The Genome Analysis Toolkit: a MapReduce framework for analyzing next-generation DNA sequencing data. Genome Res. 2010 Sep;20(9):1297–303.

[11] Kim S, Scheffler K, Halpern AL, et al. Strelka2: fast and accurate calling of germline and somatic variants. Nat Methods. 2018 Aug;15(8):591–594.

[12] Chen X, Schulz-Trieglaff O, Shaw R, et al. Manta: rapid detection of structural variants and indels for germline and cancer sequencing applications. Bioinformatics. 2016 Apr 15;32(8):1220-2.

[13] McLaren W, Gil L, Hunt SE, et al. The Ensembl Variant Effect Predictor. Genome Biol. 2016 Jun 6;17(1):122.

[14] Hagemann IS, Devarakonda S, Lockwood CM, et al. Clinical next-generation sequencing in patients with non-small cell lung cancer. Cancer. 2015 Feb 15;121(4):631–9.

[15] Simons M, Ramaekers B, Peeters A, et al. Observed versus modelled lifetime overall survival of targeted therapies and immunotherapies for advanced non-small cell lung cancer patients - A systematic review. Crit Rev Oncol Hematol. 2020 Sep;153:103035.

[16] Steuten L, Goulart B, Meropol NJ, et al. Cost Effectiveness of Multigene Panel Sequencing for Patients With Advanced Non-Small-Cell Lung Cancer. JCO Clin Cancer Inform. 2019 Jun;3:1–10.

[17] Chan TA, Yarchoan M, Jaffee E, et al. Development of tumor mutation burden as an immunotherapy biomarker: utility for the oncology clinic. Ann Oncol. 2019 Jan 1;30(1):44–56.

